# High-Frequency Activity for Language Mapping during Stereo-EEG: Comparison with Direct Cortical Stimulation

**DOI:** 10.64898/2026.04.30.26352093

**Authors:** Parveen Sagar, Matthew Hudson, Thanomporn Wittayacharoenpong, Emily Cockle, Alissandra McIlroy, Jacob Bunyamin, Joshua Laing, Matthew Gutman, Martin Hunn, Patrick Kwan, Terence J. O’Brien, Genevieve Rayner, Andrew Neal

## Abstract

**Objective:** Direct cortical stimulation (DCS) is the gold standard for language mapping during SEEG but is prone to false negatives and false positives that may contribute to post-operative dysphasia or else overly conservative resections. Task-induced high-frequency activity (HFA, 30-200Hz) is an emerging functional biomarker that may augment DCS, but its clinical utility remains uncertain. We aimed to quantify HFA’s diagnostic concordance with DCS, assessing its potential as both a surrogate marker and a screening tool.

**Methods:** In this single-centre prospective study, 23 adults undergoing SEEG completed language mapping with DCS and HFA. HFA was mapped using auditory and visual naming tasks (ANT/VNT), quantified via Morlet wavelet transforms with baseline-normalised z-scores. DCS-positive channels were those where 50Hz stimulation elicited language disruption. HFA distribution was examined independently of DCS. HFA-DCS concordance was assessed for individual and combined (ANT+VNT; maximal HFA across tasks) conditions at channel and sublobar levels across two thresholds: a specificity-optimized ‘stringent’ threshold (Z>0.8) to examine HFA as a surrogate for DCS, and a sensitivity-optimized ‘permissive’ threshold (Z>0.3) to evaluate its potential as a screening tool.

**Results:** Twelve (52%) participants were female, and 17 (74%) were MRI-negative. HFA patterns differed by task: VNT produced greater HFA magnitude in the dominant frontal lobe (p=0.0498), while ANT produced greater magnitude and activation rate in the non-dominant temporal lobe (p=0.015; p=0.0189), highest in the non-dominant superior temporal gyrus. In the combined condition, concordance with DCS was low at the stringent threshold (channel-wise sensitivity/specificity=0.24/0.88; region-wise=0.43/0.77). Sensitivity improved at the permissive threshold (channel-wise 0.56, NPV=0.96), with region-wise sensitivity of 0.75, specificity=0.45, and NPV=0.94.

**Significance:** Region-level HFA at a permissive threshold is useful for identifying language-negative regions and prioritising DCS testing. Poor concordance at a stringent threshold suggests HFA and DCS index distinct functional properties and are not interchangeable. Anatomically plausible HFA localisation supports the need for further multimodal validation to clarify its role in presurgical mapping.

**Key Points:**

- HFA and DCS show threshold- and scale-dependent diagnostic concordance for language mapping during SEEG
- Sensitivity-optimized sublobar HFA shows high negative predictive value and moderate sensitivity for DCS-positive language sites
- These metrics support sublobar HFA as a screening tool to exclude non-eloquent regions and streamline DCS language mapping
- Specificity-optimized HFA concords poorly with DCS, indicating these markers index distinct properties and are not interchangeable
- Combined HFA/DCS profiles may help stratify surgical risk: HFA-/DCS-regions as low risk, while HFA+/DCS+ sites denote high risk

## INTRODUCTION

Mapping language networks during SEEG exploration for drug-resistant focal epilepsy is essential to delineate so-called ‘eloquent cortex’ that is fundamental to speech and language function, so that subsequent resection or ablation can be tailored to minimize postoperative deficits. Direct cortical stimulation (DCS), the clinical standard, creates ‘pseudolesions’ via focal electrical current to transiently disrupt the network, inferring a region’s necessity for language. However, postoperative deficits occur despite preservation of all DCS-negative (DCS-) cortex, suggesting sensitivity limitations^1,2^. Specificity has also been questioned, as DCS may produce distant effects via current spread along white matter pathways or through highly connected nodes^3^; consequently not all DCS-positive (DCS+) sites result in postoperative deficits when resected^4^ or ablated^5^. DCS is also resource-intensive and may be limited by fatigue, after discharges, or provoked seizures, limiting completeness of testing and thereby contributing to false negatives. Consequently, there is a clinical need for complementary biomarkers to improve mapping performance.

Task-induced high-frequency activity (HFA; 30-200Hz) has been proposed as a functional mapping biomarker for language in the iEEG setting^6,7^. This activation-based approach, conceptually analogous to fMRI, involves administering language tasks during SEEG and processing the recorded signals to detect highly localized spectral power changes, from which a functional map can be inferred^8^. Compared to DCS, HFA offers rapid acquisition, the ability to assess all recorded sites without additional intervention, and reduced patient risk from non-habitual seizures and fatigue. Clinically, HFA is proposed both as an independent language mapping tool providing functional risk information, and as a screening tool to improve DCS efficiency.

Integration of HFA into clinical workflows requires multimodal validation, including correlation with postoperative outcomes and established mapping methodologies such as DCS and fMRI. Early outcome-linked studies in iEEG provide evidence for the functional importance of HFA-positive cortex^9-11^. Validation against DCS is particularly critical; although DCS is imperfect, it remains the clinical gold-standard, as resection of DCS+ sites frequently correlates with postoperative language deficits^12,13^. However, SEEG comparisons are sparse and results are mixed; recent studies report HFA sensitivity to DCS+ as low as 8.9% or no better than chance^14,15^.

These reported poor concordance rates may be driven by specific methodological choices rather than a true lack of overlap. Most previous studies have relied on stringent inferential statistics to ascertain HFA activation; while this prioritizes the reduction of false positives, it often leads to an underestimation of functionally engaged cortex. Additionally, previous analyses have focused on the contact-level comparisons, which may reduce concordance by failing to account for the broader connected regions recruited by stimulation.

These same analytic constraints also limit HFA’s utility as a screening tool to streamline DCS testing—a use-case proposed for nearly two decades. While SEEG and ECoG studies consistently report a high negative predictive value (NPV) for DCS+, suggesting HFA could efficiently exclude non-eloquent regions and reduce testing burden^7,14,16^, its low sensitivity poses an unacceptable risk of missing DCS-defined eloquent cortex.

Our primary objective was to characterize the spatial correspondence between HFA and DCS by evaluating whether addressing the methodological limitations of prior research—specifically spatial scale and activation thresholds—could improve concordance and thereby provide support for eventual integration of HFA into clinical workflows.

First, we characterised the distribution of HFA to better understand its functional relevance across canonical language regions. Then we assessed the concordance of HFA with DCS at two spatial scales: (1) channel-level (SEEG bipolar pairs) and (2) sublobar regional-level. We hypothesized that sublobar-regional analysis would improve concordance by capturing the broader functional patterns that may be recruited by stimulation.

In parallel, we evaluated whether empirically varying the quantitative thresholds for defining HFA ‘activation’ would optimize its utility for distinct clinical goals. Specifically, we examined the performance of a stringent threshold (Z > 0.8) to examine HFA as a potential surrogate for DCS-defined language cortex, and a permissive threshold (Z > 0.3) to maximize sensitivity and NPV for screening regions unlikely to be DCS+.

## METHODS

### Patient selection and the population studied

In this prospective study (Alfred Hospital, Australia, 2023–2024), 23 of 26 consecutive SEEG patients were recruited (inclusion: age >18, sufficient English/cognitive capacity). Three were excluded post-recruitment for inability to follow instructions (n=2) or tolerate testing (n=1). All underwent standard pre-implantation investigations (MRI, FDG-PET, fMRI, scalp EEG, neuropsychology).

Ethics approval was granted by the Alfred Health Research and Ethics Committee.

### SEEG study

SEEG implantation was guided solely by clinical need. Recordings were obtained using multi-contact intracerebral electrodes (8–18 contacts; Dixi Medical). Video-SEEG (Compumedics, Australia) was recorded with a 2000 Hz sampling rate and 5–1000 Hz bandpass filter. Bipolar derivations, referred to here as ‘channels’, were used for both visual inspection and signal analysis. Patients underwent a planned ASM withdrawal with the aim of capturing habitual seizures.

### Direct cortical stimulation process

DCS was performed 4–5 days post-implantation using our standard local protocol for language mapping, as detailed in Cockle et al.^5^. In brief, high frequency bipolar stimulation (50 Hz, 0.5ms pulse width, 5s duration, 0.5–3mA) was used. Current was incrementally increased until one of the following occurred: (i) seizure, (ii) a clinical response (positive or negative), (iii) afterdischarges or (iv) the 3mA threshold was reached. Channels were classified as DCS+ if stimulation elicited reproducible language disruption, as determined by mutual agreement between a neurologist and neuropsychologist. Language tasks included visual naming, auditory naming, spontaneous speech, counting, and reading. Because task selection did not always permit differentiation between motor speech and expressive or receptive deficits, all such disturbances were combined pragmatically as “language deficits”. For the purpose of analysis, trials involving afterdischarges and seizures were excluded, as these precluded the reliable assessment of primary language disruption.

### High frequency activity mapping process

#### Language tasks used for HFA mapping

We adapted the Auditory Naming Test^17^ (ANT) and Visual Naming Test^18^ (VNT) as they both have established validity for clinical assessment of language processes. These tasks recruit partly distinct neural systems^17^, increasing identification of language-relevant cortex, and are routinely used in pre-implantation assessment and DCS. To maximise signal-to-noise ratio, tasks followed a structured, short-epoch, repeating format.

#### HFA mapping process

HFA mapping was typically performed on day 5-6 of the admission, usually one day after cortical stimulation and always prior to radiofrequency thermocoagulation (RF-THC). Routine SEEG monitoring continued in parallel, with sleep deprivation and medication adjustments guided by clinical needs. Testing was conducted while patients rested in a standard hospital bed within a shared 4-bed bay.

Visual stimuli were presented using PowerPoint (Microsoft) slideshows on a laptop, which advanced automatically to maintain precise timing. Fig. 1 illustrates the setup used for both tasks.

**Figure 1:**
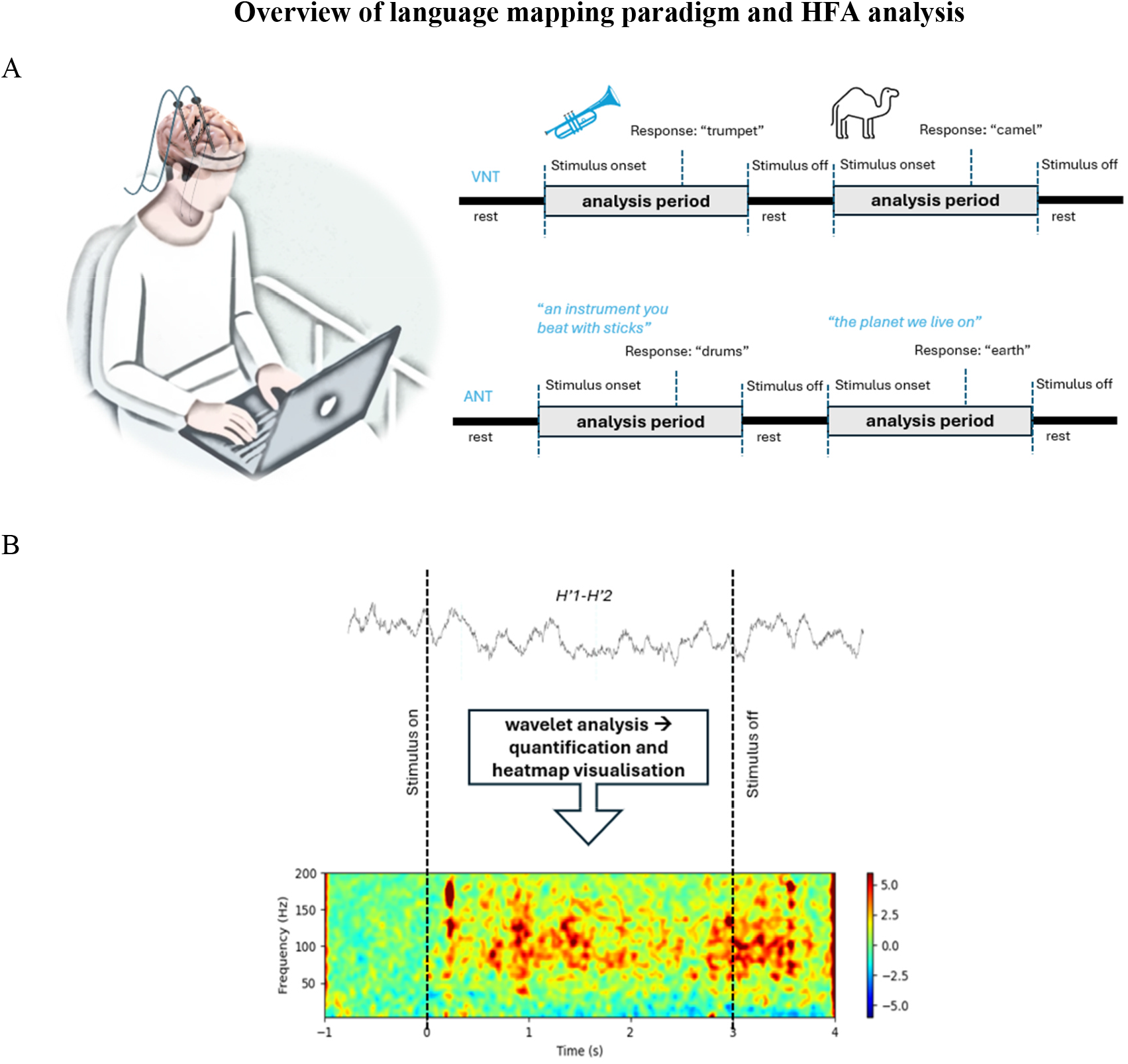
Overview of language mapping paradigm and HFA analysis. (A) Schematic of the VNT and ANT, illustrating task structure and timing. During VNT, patients named images presented for 3 seconds; during ANT, they responded to pre-recorded auditory prompts (∼5 seconds). Each stimulus was followed by a rest period to allow recovery. (B) Example of HFA analysis. Raw SEEG signals were transformed using Morlet wavelets to compute time– frequency representations. Spectral power was baseline-normalized using z-scores, averaged across time and frequency bands of interest, and visualized as heatmaps.

The VNT comprised 30 images of single objects presented for 3s each, interleaved with 2s rest periods (total duration 3 min)^19^. White noise was delivered via passive noise-cancelling headphones to minimise environmental noise.

The ANT comprised 50 pre-recorded auditory stimuli (∼5s each) followed by 2s rest periods (total duration 6 min). White noise was not used to preserve auditory stimulus clarity. The tasks were administered in a fixed order, and patients were instructed to wait until the listening period ended to respond.

Stimulus onset and offset were time-stamped on the SEEG recording using StimTracker hardware (Cedrus), via photodiode-detected TTL pulses to synchronise task events with electrophysiological data.

#### HFA signal processing

At completion of tasks, the corresponding segment of raw SEEG was extracted and analysed using a custom Python script (key library: MNE). Epochs of interest were defined relative to baseline: −0.5 to 0s for baseline, and 0 to 3s (VNT) or 0 to 5s (ANT) for task execution.

Event-related spectral perturbations (ERSPs) were computed using Morlet wavelet transforms, with spectral power baseline-normalised on a z-scale. High-frequency activity (30–200 Hz) was quantified by averaging task-related power changes within the predefined task window for each channel. Channels exceeding z-score thresholds of 0.8 or 0.3 were classified as HFA-positive (HFA+) and compared with DCS results for concordance analysis. Time–frequency heatmaps were generated for qualitative review using MNE plotting functions.

#### Data cleanup

Because analyses included epileptogenic cortex, artefact rejection was essential to avoid spuriously elevated HFA. Intracerebral contamination from interictal epileptiform discharges (IEDs) was addressed using a two-step expert review process: first, task epochs were visually screened to remove epochs containing widespread IED contamination; second, time–frequency representations were reviewed to identify channels with persistent IED-related broadband contamination, which were excluded from further analysis. To minimise extracerebral artefact, contacts identified as non-cerebral (e.g. dura or bone) and their immediate neighbours were removed. Bipolar pairs traversing white matter or multiple sublobar regions were also excluded.

### Characterisation and regional distribution of HFA

We characterized the distribution of HFA and assessed for significant differences across hemispheres, lobes and sublobar regions. This was conducted independently of DCS status to identify regions that consistently showed HFA during task execution.

HFA activation was defined using the ‘stringent threshold’ (Z>0.8) (described below), identifying channels with robust task-induced signal changes. Activation rates were calculated as the proportion of channels exceeding this threshold within each region. To ensure representative sampling, analysis was restricted to sublobar areas containing ≥4 bipolar channels. Activation rate and magnitude of HFA (mean Z-scores) were compared on hemispheric, lobar and sublobar levels.

Channel-level hemispheric differences were assessed using Mann–Whitney U tests (magnitude) and Fisher’s exact tests (activation rate). Sublobar-regional activation rates were compared using paired Wilcoxon signed-rank tests, and region-wise hemispheric differences in magnitude and activation rate were assessed using Mann–Whitney U and Fisher’s exact tests, respectively. Lobar analyses were conducted similarly. False discovery rate (FDR) correction was applied where multiple comparisons were performed.

### Examining Concordance Between HFA and DCS

To evaluate the clinical relevance of HFA for language mapping, we compared it to DCS across two levels of analysis: (i) a channel-wise analysis, comparing HFA to DCS status at the level of individual channels; and (ii) a sublobar region-wise analysis. Concordance was assessed for the VNT and ANT tasks separately, as well as together (ANT+VN), defined by the maximum value across both tasks. Only DCS tested channels were included, as unstimulated sites could not be reliably classified as DCS-negative - consistent with prior studies ^7,14,20^.

Confusion matrices were constructed to calculate sensitivity, specificity, positive predictive value (PPV), and negative predictive value (NPV) for HFA in predicting DCS status, with 95% confidence intervals estimated using the Wilson score method. All analyses were performed using Python (NumPy, SciPy, and Matplotlib).

#### Threshold selection for defining HFA activation

Given the absence of a universally accepted thresholding framework for defining HFA activation (as discussed in our recent narrative review^8^), thresholds were empirically calibrated within this cohort to align with specific clinical objectives rather than applying a fixed statistical criterion.

First, to assess HFA as a marker of DCS-defined language-positive cortex, a ‘stringent threshold’ of Z = 0.8 was applied. This value corresponds to the 80th percentile of the regional HFA distribution within this cohort. This threshold prioritises specificity, consistent with the intended role of HFA as a confirmatory marker of DCS-defined eloquent cortex.

Second, to evaluate the utility of HFA as a screening tool, a ‘permissive threshold’ of Z = 0.3 was applied. This corresponds to the 50^th^ percentile of the overall HFA distribution in our cohort. This value represents the minimum task-related power change considered physiologically significant, set to maximize sensitivity while remaining above the empirical noise floor.

### Region-wise Concordance between HFA and DCS

#### Anatomical Labelling Framework for Region-Wise Analysis

To enable regional analysis, sublobar regions of interest that are meaningful in the SEEG context were defined using a bespoke hierarchical naming convention informed by SEEG labelling practice. This system used a three-level structure: (lobe.sublobarregion.[subdivision]; see Supplementary Table 1).

For each patient, contact localization was performed via co-registration of pre-operative MRI and post-implantation CT, followed by manual assignment of anatomical labels to each contact. Language dominance was determined on a per-patient basis using pre-operative language fMRI, or if unavailable, DCS mapping.

#### Region-wise classification criteria

In each patient, channels sharing the same sublobar label were grouped. Unlike the descriptive analysis, which used a ⩾4 channel threshold for statistical stability, this concordance analysis included all regions containing ⩾1 DCS-tested channel to maximise anatomical coverage.

HFA-positive regions were defined as those with at least one channel exceeding the respective z-score threshold (stringent or permissive), depending on the objective. DCS-positive regions were those in which ⩾1 channel elicited a language-related response during stimulation.

## RESULTS

### Clinical characteristics

Table 1 presents clinical characteristics (details in Supp. Table 2). After excluding IED and movement degraded channels (n=76), 1955 remained. Channels with stimulation-elicited afterdischarges (n=117) were also excluded.

**Table 1:** Demographic and clinical characteristics of enrolled participants (N=23)

| Variable | Value |
| --- | --- |
| **Demographics** |  |
| Mean age at SEEG, years (range) | 34.7 (20-50) |
| Male sex (n, %) | 12 (52%) |
| Left-handedness (n, %) | 4 (17%) |
| **Epilepsy Characteristics** |  |
| Mean age of epilepsy onset, years (range) | 17.5 (0-40) |
| Median number of ASM at time of SEEG (range) | 3 (2-4) |
| Epileptogenic MRI Lesion (n, %) | 6 (26%) |
| Focal FDG-PET hypometabolism (n, %) | 20 (87%) |
| EZ in dominant hemisphere (n, %) | 15 (65%) |
| **Sampling Laterality** |  |
| Left hemisphere only (n, %) | 7 (30%) |
| Right hemisphere only (n, %) | 2 (9%) |
| Bilateral implantation (n, %) | 14 (61%) |
| **Post SEEG EZ formulation** |  |
| Frontal (n,%) | 3 (13%) |
| Temporal (n,%) | 14 (61%) |
| Parietal (n,%) | 4 (17%) |
| Occipital (n,%) | 1 (4.5%) |
| Insular (n,%) | 1 (4.5%) |
| Abbreviations: ASM, anti-seizure medication; EZ, epileptogenic zone |  |

Of the remaining channels, 624 (38%) were stimulated with 50Hz stimulation for language mapping and were included in the final analysis. DCS+, defined as the presence of a language disruption during stimulation was observed at 45 channels (6.4%).

### Location of DCS sites

The anatomical distribution of DCS-tested and DCS+ sites is illustrated in Fig. 2. The majority of stimulations were located in the temporal lobes (54.3%), and the dominant temporal and frontal lobes yielded the highest rates of DCS+ (12.8% and 12.2%, respectively).

**Figure 2:**
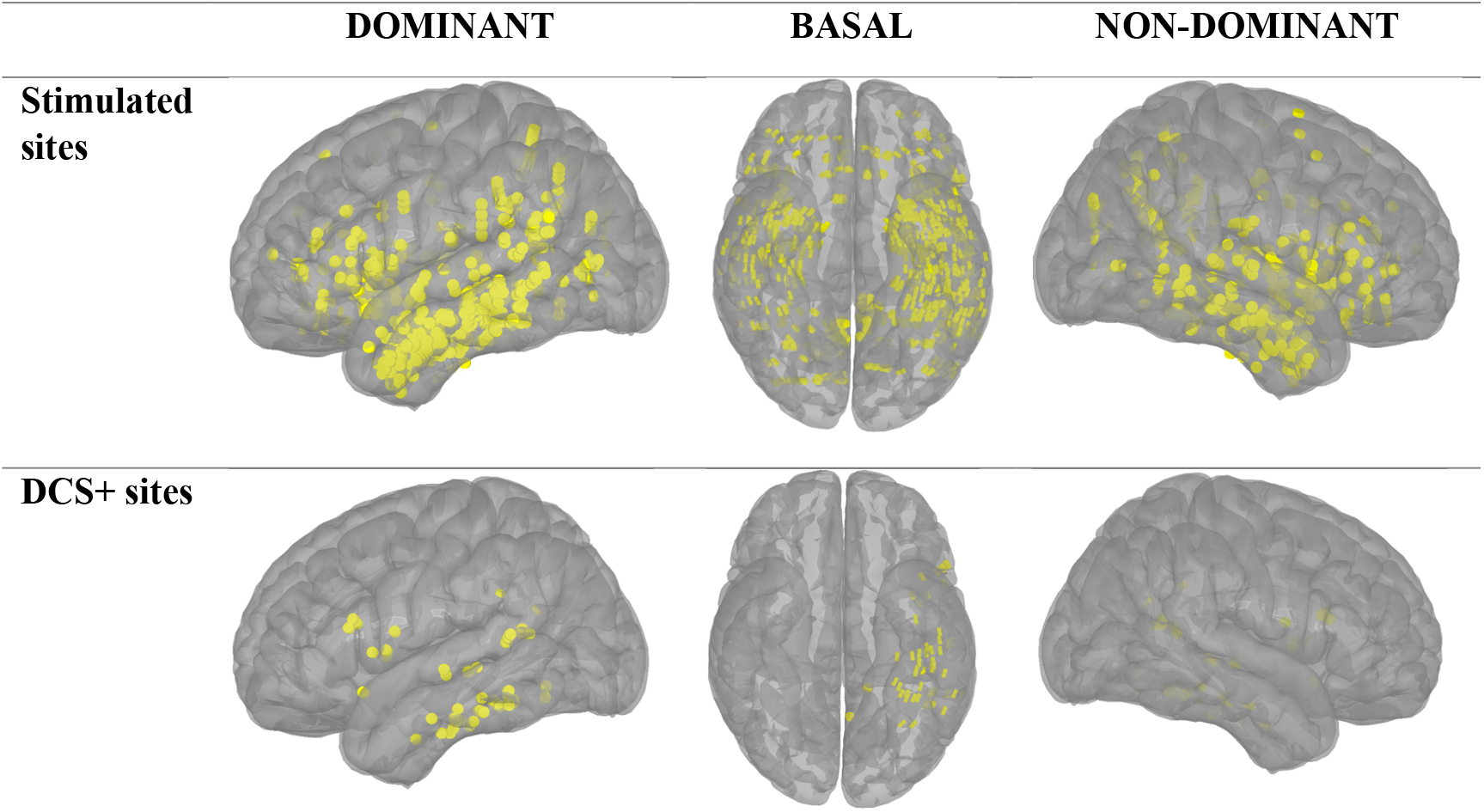
The distribution of DCS tested and DCS+ sites across 23 patients. Visualisation created in Brainstorm.

### Location of HFA positive sites

HFA activation was distributed across a wide range of cortical regions, with spatial patterns differing by task (Fig. 3). For VNT, channel-wise activation rates did not differ between hemispheres (Table 2). Likewise, no hemispheric differences were observed in sublobar-regional activation-rate comparisons. When analysed at the lobar level, mean HFA magnitude differed between hemispheres in the frontal (FDR-corrected p = 0.0498) and parietal lobes (FDR-corrected p = 0.0146). The dominant hemisphere demonstrated higher magnitude in the frontal lobe, whereas the non-dominant hemisphere showed higher magnitude in the parietal lobe.

**Figure 3:**
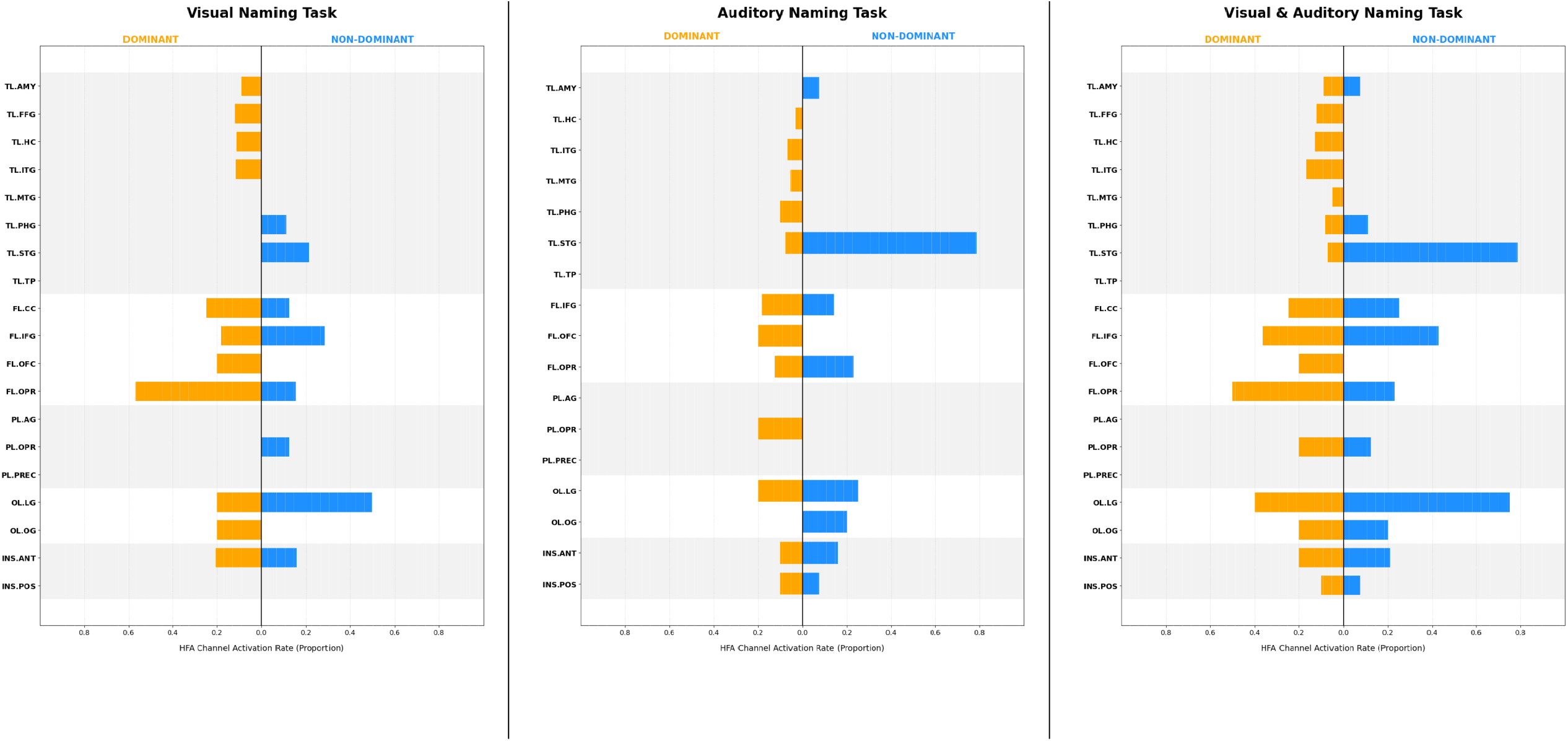
HFA activation rates (Z > 0.8) across sublobar brain regions, stratified by language task. For ease of interpretation, only the second level of the hierarchical region labels is displayed (e.g., TL.FFG). Each bar represents the proportion of channels exceeding Z>0.8 threshold within that region. Detailed regional activation proportions for each task and hemisphere are provided in Supplementary Table 3. Abbreviations: **TL (Temporal Lobe):** TP, temporal pole; STG, superior temporal gyrus; PHG, parahippocampal gyrus; MTG, middle temporal gyrus; ITG, inferior temporal gyrus; HC, hippocampus; FFG, fusiform gyrus; AMY, amygdala. **FL (Frontal Lobe):** OPR, operculum; OFC, orbitofrontal cortex; IFG, inferior frontal gyrus; CC, cingulate cortex. **PL (Parietal Lobe):** PREC, precuneus; AG, angular gyrus. **OL (Occipital Lobe):** OG, occipital gyri; LG, lingual gyrus.

**Table 2:** HFA Activation Rates at the Hemisphere level.

| TASK | HEMISPHERE | CHANNELS<br>(N) | HFA<br>ACTIVE<br>(N, %) | MEAN<br>Z-SCORE | ACTIVATION<br>RATE |
| --- | --- | --- | --- | --- | --- |
| VNT | Non-Dominant | 227 | 18 (7.9%) | 0.19 | 0.08 |
|  | Dominant | 366 | 32 (8.7%) | 0.17 | 0.09 |
| ANT | Non-Dominant | 209 | 24 (11.5%) | 0.24*<br>($p=0.05$) | 0.11*<br>($p=0.015$) |
|  | Dominant | 354 | 20 (5.6%) | 0.09 | 0.06 |
| VNT &<br>ANT | Non-Dominant | 231 | 34 (14.7%) | 0.37 | 0.15 |
|  | Dominant | 392 | 46 (11.7%) | 0.27 | 0.12 |

Summary statistics for all recorded bipolar SEEG channels. HFA activation was tested at a Z-score threshold of 0.8 relative to the pre-stimulus baseline. Asterisks indicate significant hemispheric differences ( $p<0.05$ ).

In contrast, ANT demonstrated a non-dominant hemispheric bias across both activation rate (p=0.015) and HFA magnitude (p=0.041) (Table 2). Moreover, the non-dominant temporal lobe demonstrated significantly greater HFA magnitude (FDR-corrected p=0.0397) and a higher activation rate (FDR-corrected p=0.0189) compared with the dominant hemisphere (Supp. table 2). At the sublobar level, this asymmetry was focal, with the superior temporal gyrus (TL.STG) showing a higher HFA magnitude (0.53 vs 0.0013, FDR-corrected p=0.03) and activation rate in the non-dominant hemisphere, though this did not survive FDR correction (79% [11/14] vs. 8% [3/27]; FDR-corrected p = 0.6488).

When data from both tasks were combined (VN+ANT), no significant hemispheric, or lobar differences were seen, though the superior temporal gyrus (TL.STG) once again demonstrated greater HFA magnitude and activation rates (11/14 vs 2/29, FDR-corrected p=0.0001) non-dominantly.

### Channel-wise diagnostic performance of HFA activation

When examined at the channel level, both the VNT and ANT paradigms showed poor concordance with DCS using both the stringent and permissive thresholds (Fig 4A; Supp Table 4 and 5). Across all conditions, sensitivity and PPV were low, with slightly higher sensitivity seen for VNT (0.18) compared to ANT (0.09). The combined condition (ANT+VN) yielded a marginally higher sensitivity (0.24). Specificity and negative predictive value (NPV) were consistently high across all conditions, reflecting the rarity of DCS-positive channels. Balanced accuracy (BA) remained below 0.60 for all conditions.

**Figure 4:**
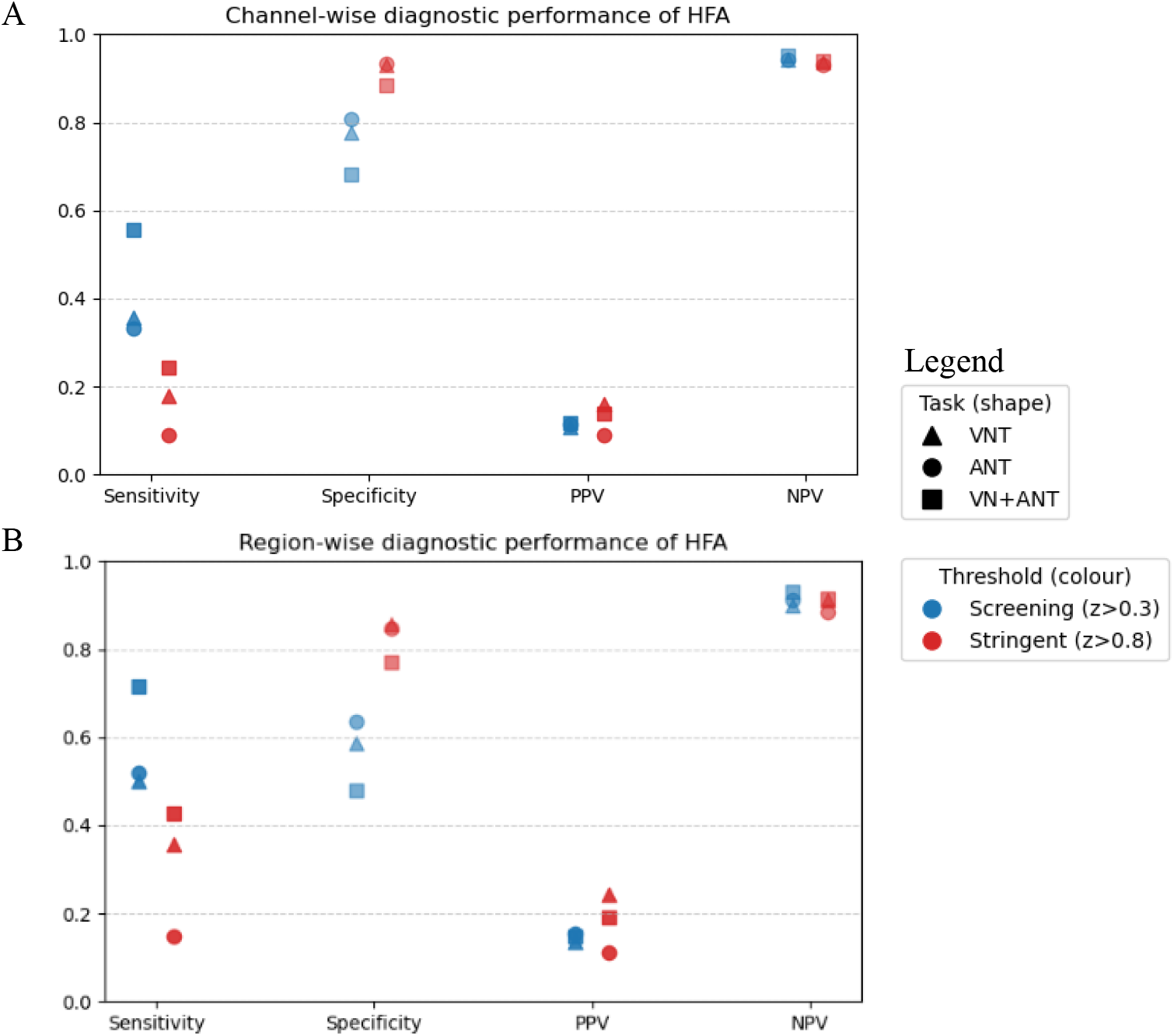
Channel- and region-wise diagnostic performance of task-induced HFA relative to DCS. (A) Channel-level and (B) sublobar region-level sensitivity, specificity, PPV, and NPV for ANT, VNT, and combined (ANT+VNT) conditions. Metrics are shown for the permissive threshold (z > 0.3; blue) and stringent threshold (z > 0.8; red).

### Region-wise diagnostic performance

When examined at the regional level, both the VNT and ANT paradigms showed improved diagnostic performance compared to the channel-level analysis. At the stringent threshold (Z > 0.8; Figure 4B; Supp Table 6), specificity increased (0.77) but at the cost of reduced sensitivity (0.43), with BA remaining comparable to the permissive threshold (0.59 vs. 0.60). At the permissive threshold (Z > 0.3; Figure 4B; Supp table 7), the combined ANT+VNT condition demonstrated high sensitivity (0.75) and NPV (0.94), but specificity (0.45) and positive predictive value (0.13) remained low.

### False negative regions at the stringent threshold

For the combined ANT+VNT condition, 16 DCS+ regions were classified as HFA-negative. These false negatives were most frequently located in the dominant fusiform gyrus (*n* = 5), followed by the posterior superior temporal gyrus (*n* = 2), inferior temporal gyrus (*n* = 2), and posterior cingulate cortex (*n* = 2). Additional sites included the middle temporal gyrus (n=1), anterior fusiform gyrus (n=1), and parahippocampal gyrus (n=1).

## DISCUSSION

Our findings show that the clinical value of HFA for language mapping is dependent on the analytic framework. By addressing the methodological limitations of prior research —specifically spatial scale and activation thresholds—we show that HFA can serve as an effective screening tool to streamline DCS but is not a direct surrogate for DCS-defined necessity. Specifically, sublobar-aggregated HFA analysed with a permissive, sensitivity-optimized threshold (Z>0.3) offers a high NPV and moderate sensitivity. In contrast, concordance was poor at the stringent specificity-optimized threshold (Z>0.8), suggesting that HFA and DCS index distinct but complementary functional properties.

### Screening potential of HFA for DCS sites

Sublobar-aggregated HFA analyzed with a sensitivity-optimized threshold (Z>0.3) yielded better diagnostic performance for screening than the channel-level analysis. Combining ANT and VNT conditions resulted in a sensitivity of 0.75 and an NPV of 0.94. These metrics indicate that HFA-negative regions are highly unlikely to be DCS-positive for language. While specificity and PPV were low, this trade-off is acceptable for a screening tool designed to rule out eloquent cortex and streamline clinical workflows. Conversely, channel-wise HFA performed poorly (sensitivity 0.56), reinforcing that contact-level analysis is unsuitable for screening.

Several methodological choices, designed to overcome limitations of prior studies, explain this superior screening performance compared with the low sensitivity (e.g., 8.9%) reported in prior research^14,15^. First, sublobar aggregation likely improves concordance, as DCS effects may not be ultra focal^3^. Focusing on peak regional activation may therefore better approximate the connected cortex recruited by stimulation.

Secondly, the application of a permissive activation threshold ensured that subtle but functionally relevant cortex was not excluded^7^. Stringent inferential statistics used in earlier studies prioritize the reduction of false positives, which often leads to an underestimation of functionally engaged cortex.

Thirdly, our broader analysis window (30–200Hz) also likely improved performance; lower gamma frequencies (30–50Hz) better detect DCS+ sites. Prior studies often utilize narrower high-gamma bands (e.g., 70–150 Hz), which are highly specific but less sensitive and thus may miss broader activations^21^. Notably, frequency selection varies widely across prior studies, with inconsistent lower and upper bounds^7,14,20,22^, while the optimal range for predicting DCS remains unknown^23^. Lastly, combining language conditions also improves sensitivity by extending coverage across language-related regions engaged by each task^17^.

### HFA as a Surrogate for DCS-defined Language Cortex

We found that HFA has poor concordance with DCS at our stringent threshold, regardless of whether analysed at the channel or regional level. This indicates that, when evaluated as a potential surrogate for DCS-defined necessity, HFA performs poorly. Sensitivity remained low across task conditions, with 16 DCS+ regions failing to show HFA at the stringent threshold. While NPV was consistently high (reflecting the rarity of DCS+ sites), balanced accuracy was only marginally above chance. These findings confirm that HFA should not be interpreted as a surrogate for DCS-defined language cortex.

Our findings align with prior SEEG studies; one reported 8.9% sensitivity using a contact-wise approach and another demonstrated low concordance despite utilising a regional framework^14,15^. In contrast, ECoG studies have reported higher sensitivity, with a wide range from 0.22 to 0.99, though pooled sensitivity (0.61) remains moderate overall^16^. However, key methodological differences limit direct comparison with SEEG. Subdural grids used in ECoG typically sample cortex contiguously and are often placed over presumed language areas, increasing the pre-test probability of detecting language-positive sites. In contrast, SEEG sampling is sparser and hypothesis-driven, leading to a greater proportion of truly negative sites. This difference inflates NPV and deflates sensitivity in the SEEG context and underscores the lack of generalizability of ECoG-derived performance metrics.

#### Reasons for poor concordance at the stringent threshold

The observed dissociation—where regional HFA excels as a screening tool yet fails as a surrogate for DCS-defined necessity—arises from fundamental differences between the two modalities, which our data illustrate.

First, DCS is an inactivation technique that creates transient ‘pseudolesions’ anticipating resection or ablation and probes functional necessity. In contrast, HFA is an activation-based method that reflects cortical engagement^7,24^. Given this mechanistic difference, low concordance is not wholly unexpected. Still, it may be assumed that task-critical regions will show robust activation identifiable at the stringent threshold. However, this may not hold if certain regions act as connector hubs—critical for task execution but not associated with strong local HFA^25^. In our cohort, several DCS+ regions were HFA-negative at the stringent threshold, particularly within ventral and posterior temporal areas. This suggests that language-processing in these regions may rely on integrative or relay functions that produce only modest or distributed increases in spectral power, explaining its frequent HFA-negativity in our study.

Second, unlike DCS, HFA lacks a binary outcome. It is unclear what magnitude of HFA signifies meaningful cortical engagement, so thresholds were set empirically^7,8^. At our stringent threshold sensitivity was low, confirming that a high magnitude of local HFA is not a reliable proxy for a region’s indispensability. Similar thresholding issues are documented in fMRI validation studies^26,27^.

Third, DCS can produce network-level effects by disrupting remote but connected regions^3^, while HFA typically reflects localized processing on a millimetre scale^23,28-30^. This spatial mismatch complicates direct comparison. To address this, we used regional maxima, which may better approximate the broader cortical zone impacted by stimulation.

### Biological Plausibility of HFA distribution

Task-induced HFA was observed in canonical language regions across the two testing paradigms. Lateralization patterns varied by task: the VNT elicited significantly greater activation in the dominant frontal lobe, consistent with the role of the anterior language networks in motor speech production as well as lexical generation and retrieval^31,32^, whereas the ANT evoked significantly greater non-dominant activation, with the strongest effect seen in the STG—where activity was both prominent and lateralized. While this pattern aligns generally with the STG’s established role in auditory lexical access and phonological decoding ^33^, the strong non-dominant lateralisation is unexpected. In our cohort, we cannot definitively distinguish between potential explanations such as language reorganisation in patients with dominant-hemisphere epilepsy or task-specific recruitment of the non-dominant STG for prosodic or spectral analysis. Individual-level analysis was underpowered to test these hypotheses. Further studies are needed to determine whether this pattern is common in auditory language tasks or specific to this cohort. Together, these findings suggest that even in the absence of strong concordance with DCS, HFA captures task-relevant cortical engagement across the distributed language network.

### Clinical integration of HFA in presurgical language mapping

Our findings support a complementary framework in which HFA and DCS provide distinct but clinically relevant information for language mapping. Outside the epileptogenic zone, HFA can screen for areas unlikely to be DCS+, prioritizing stimulation targets. In practice, screening with regional-HFA may be most effective when guided by pre-test clinical judgment—using HFA-negativity to support the exclusion of regions already considered unlikely to be language-critical. However, within the EZ or regions being considered for resection or ablation, HFA alone cannot exclude language function; DCS remains essential.

Within this framework, combined HFA and DCS profiles may help stratify language risk. HFA–/DCS– regions likely represent low-risk tissue, whereas HFA+/DCS+ regions denote high-risk eloquent cortex. Discordant profiles (e.g., HFA+/DCS–) may reflect functionally engaged but stimulation-negative cortex, where resection could still carry risk. Consistent with this, outcome-linked studies in both ECoG and SEEG have demonstrated that resection of HFA+, DCS– cortex may be associated with postoperative language decline^9,10^, suggesting that HFA can capture functionally meaningful cortex not identified by stimulation alone.

Accordingly, HFA is best conceptualised not as a surrogate for DCS-defined language-essential cortex, but as a complementary signal within a multimodal presurgical mapping framework that warrants continued outcome linked validation.

## Limitations and future directions

This single-centre study had a modest sample size and variable sublobar sampling inherent to SEEG. Therefore, sublobar coverage was incomplete, limiting generalisability and full network characterisation. Larger multi-centre cohorts will be important to improve anatomical coverage and statistical power.

HFA artefact rejection was performed via expert review and is subjective and time intensive. Retention of ambiguous signals may inflate activation rates, or vice versa. Automated approaches, such as machine learning-based artefact detection, may improve reproducibility and reduce rater dependence. Our analysis focused on raw HFA power changes only. Other spectral features may improve predictive performance. Finally, thresholds were set empirically within this cohort and may not generalise to other centres; outcome-based optimisation in independent prospective cohorts will be necessary before clinical implementation.

DCS itself is an imperfect comparator. As an inactivation method, it can produce network-level effects, leading to false positives, or false negatives may result from subthreshold stimulation, afterdischarges, or seizures. Limited concordance between HFA and DCS should therefore not be interpreted as definitive evidence against HFA’s construct validity.

Future work should prioritise outcome-linked validation. Correlating HFA-positive regions with postoperative language deficits following resection or ablation will better define its clinical utility. Prospective studies should also assess whether HFA-guided screening can reduce DCS duration and procedural risk.

## Conclusion

Task-induced region-level HFA may be used in SEEG language mapping as a screening tool to exclude non-eloquent cortex from DCS testing. However, overall poor concordance at the stringent threshold indicates that HFA should not be interpreted as a surrogate for DCS, but as a complementary tool within multimodal presurgical mapping frameworks. Further multimodal and outcome-based validation will be essential to define its role within the presurgical mapping toolkit.

## Data Availability

All data produced in the present study are available upon reasonable request to the authors

## Acknowledgements

PS is supported by an Australian Government Research Training Program (RTP) Scholarship. The authors also gratefully acknowledge the patients who generously participated in this research, as well as the support of the wider Alfred Health SEEG team.

