## Supplementary tables for "High-Frequency Activity for Language Mapping during Stereo-EEG: Comparison with Direct Cortical Stimulation"

### Supplementary material

**Supplementary Table 1: Sublobar Region Labels Used for Region-Wise Analysis.**
Labels follow a lobe.region.subclassification format. Subclassifications were applied only when functionally relevant.

| ABBREVIATION | FULL NAME |
|---|---|
| **FL.CC** | Frontal Lobe: Cingulate Cortex |
| **FL.FP** | Frontal Lobe: Frontal Pole |
| **FL.IFG.PO** | Frontal Lobe: Inferior Frontal Gyrus: Pars Opercularis |
| **FL.IFG.PORB** | Frontal Lobe: Inferior Frontal Gyrus: Pars Orbitalis |
| **FL.IFG.PT** | Frontal Lobe: Inferior Frontal Gyrus: Pars Trianpgularis |
| **FL.MFG** | Frontal Lobe: Middle Frontal Gyrus |
| **FL.OFC** | Frontal Lobe: Orbitofrontal Cortex |
| **FL.OPR** | Frontal Lobe: Frontal Operculum |
| **FL.PRECG** | Frontal Lobe: Precentral Gyrus |
| **FL.PREM** | Frontal Lobe: Premotor Area |
| **FL.SFG** | Frontal Lobe: Superior Frontal Gyrus |
| **INS.ANT** | Insula: Anterior |
| **INS.POS** | Insula: Posterior |
| **OL.LG** | Occipital Lobe: Lingual Gyrus |
| **OL.OG** | Occipital Lobe: Occipital Gyrus |
| **OL.SUPRAC** | Occipital Lobe: Supracalcarine Area |
| **PL.AG** | Parietal Lobe: Angular Gyrus |
| **PL.CC** | Parietal Lobe: Cingulate Cortex |
| **PL.OPR** | Parietal Lobe: Parietal Operculum |
| **PL.POCG** | Parietal Lobe: Postcentral Gyrus |
| **PL.PREC** | Parietal Lobe: Precuneus |
| **PL.SMG** | Parietal Lobe: Supramarginal Gyrus |
| **PL.SPL** | Parietal Lobe: Superior Parietal Lobule |
| **TL.AMY** | Temporal Lobe: Amygdala |

| | |
|---|---|
| **TL.FFG.ANT** | Temporal Lobe: Fusiform Gyrus: Anterior |
| **TL.FFG.POS** | Temporal Lobe: Fusiform Gyrus: Posterior |
| **TL.HC** | Temporal Lobe: Hippocampus |
| **TL.HC.POS** | Temporal Lobe: Hippocampus: Posterior |
| **TL.ITG.ANT** | Temporal Lobe: Inferior Temporal Gyrus: Anterior |
| **TL.ITG.POS** | Temporal Lobe: Inferior Temporal Gyrus: Posterior |
| **TL.MTG.ANT** | Temporal Lobe: Middle Temporal Gyrus: Anterior |
| **TL.MTG.POS** | Temporal Lobe: Middle Temporal Gyrus: Posterior |
| **TL.OPR** | Temporal Lobe: Temporal Operculum |
| **TL.PHG.ANT** | Temporal Lobe: Parahippocampal Gyrus: Anterior |
| **TL.PHG.POS** | Temporal Lobe: Parahippocampal Gyrus: Posterior |
| **TL.STG.ANT** | Temporal Lobe: Superior Temporal Gyrus: Anterior |
| **TL.STG.HG** | Temporal Lobe: Superior Temporal Gyrus: Heschel's Gyrus |
| **TL.STG.POS** | Temporal Lobe: Superior Temporal Gyrus: Posterior |
| **TL.STG.PT** | Temporal Lobe: Superior Temporal Gyrus: Planum Temporale |
| **TL.TP** | Temporal Lobe: Temporal Pole |

**Supplementary Table 2: Table showing detailed demographic and clinical characteristics of enrolled patients.**

| Pt # | Age Range | Sex | Age at seizure onset | Handedness | Language lateralisation (fMRI) | SEEG implant lateralisation | Epileptogenic zone location post SEEG |
|---|---|---|---|---|---|---|---|
| 1 | 40-49 | Female | 30-39 | Right | Left | Left | left mesial temporal + insula |
| 2 | 20-29 | Male | 0-10 | Right | Left* | Bilateral | bimesial temporal + right insula |
| 3 | 20-29 | Female | 10-19 | Left | Left | Left | left inferior frontal sulcus, FCD signature |
| 4 | 30-39 | Male | 10-19 | Right | Left | Bilateral | right aMCC, FCD signature |
| 5 | 50-59 | Female | 40-49 | Left | Left | Bilateral | bimesial temporal |
| 6 | 20-29 | Male | 10-19 | Right | Right | Bilateral | diffuse left posterior quadrant |
| 7 | 30-39 | Female | 10-19 | Right | Left | Left | left inferior frontal sulcus, FCD pattern |
| 8 | 40-49 | Male | 17=0-19 | Right | Left | Bilateral | right insula + mesial temporal |
| 9 | 30-39 | Female | 0-10 | Right | Left | Bilateral | bimesial temporal |
| 10 | 30-39 | Male | 10-19 | Right | Left | Bilateral | right temporo-occipital + mesial temporal + insular |
| 11 | 40-49 | Male | 10-19 | Right | Left | Right | right mesial temporal + right STG |
| 12 | 30-39 | Male | 0-10 | Right | Left | Bilateral | right mesial temporal |
| 13 | 40-49 | Male | 0-10 | Right | Left | Bilateral | left internal parietal operculum |
| 14 | 20-29 | Male | 10-19 | Right | Left | Left | left posterior STG |

| | | | | | | | |
|---|---|---|---|---|---|---|---|
| 15 | 30-29 | Female | 20-29 | Right | Left* | Left | left anterior lateral temporal |
| 16 | 30-29 | Male | 0-10 | Left | Left | Bilateral | left mesial temporal |
| 17 | 30-39 | Female | 20-29 | Right | Left | Bilateral | left amygdala |
| 18 | 20-29 | Female | 10-19 | Right | Left | Left | left anterior fusiform gyrus |
| 19 | 40-49 | Male | 40-49 | Right | Left | Bilateral | right mesial temporal |
| 20 | 20-29 | Female | 20-29 | Left | Left | Right | right mesial temporal |
| 21 | 50-59 | Female | 20-29 | Right | Left | Bilateral | left subcentral operculum/peri-rolandic |
| 22 | 40-49 | Female | 20-29 | Right | Left* | left | left mesial temporal |
| 23 | 40-49 | Male | 30-39 | Right | Left | Bilateral | right mesial temporal |

* fMRI unavailable; lateralisation determined by direct cortical stimulation language testing.

**Supplementary Table 3: Sublobar-regional HFA activation proportions across language tasks.** Regional HFA activation proportions are displayed as ratios of active channels to the total number of channels sampled (n/N) within each sublobar region. Activation was defined using the stringent threshold of Z>0.8 relative to the pre-stimulus baseline. Data are stratified by task—Visual Naming (VNT), Auditory Naming (ANT), and the combined task condition (VNT+ANT)—and by hemispheric laterality relative to language dominance (DOM: dominant; NONDOM: non-dominant). For anatomical definitions of sublobar regions, see Supplementary Table 1.

| LOBE | SUBLOBAR LOCATION | VNT DOM | VNT NONDOM | ANT DOM | ANT NONDOM | VNT+ANT DOM | VNT+ANT NONDOM |
|---|---|---|---|---|---|---|---|
| **TL** | TL.AMY | 2/22 | 0/13 | 0/18 | 1/13 | 2/23 | 1/13 |
| | TL.FFG | 4/33 | 0/4 | 0/29 | 0/3 | 4/33 | 0/4 |
| | TL.HC | 4/35 | 0/23 | 1/32 | 0/19 | 5/39 | 0/23 |
| | TL.ITG | 2/17 | 0/4 | 1/15 | 0/5 | 3/18 | 0/6 |
| | TL.MTG | 0/34 | 0/15 | 2/39 | 0/14 | 2/40 | 0/15 |
| | TL.OPR | 0/3 | 1/2 | 0/3 | 0/2 | 0/3 | 1/2 |
| | TL.PHG | 0/24 | 1/9 | 2/20 | 0/4 | 2/25 | 1/9 |
| | TL.STG | 0/27 | 3/14 | 2/26 | 11/14 | 2/29 | 11/14 |
| | TL.TP | 0/25 | 0/19 | 0/23 | 0/15 | 0/27 | 0/19 |
| **FL** | FL.CC | 1/4 | 1/8 | 0/3 | 2/8 | 1/4 | 2/8 |
| | FL.IFG | 2/11 | 2/7 | 2/11 | 1/7 | 4/11 | 3/7 |
| | FL.MFG | 1/4 | 0/1 | 0/4 | 0/1 | 1/4 | 0/1 |
| | FL.OFC | 1/5 | 0/10 | 1/5 | 0/6 | 1/5 | 0/10 |
| | FL.OPR | 4/7 | 2/13 | 1/8 | 3/13 | 4/8 | 3/13 |
| | FL.PRECG | - | 1/4 | - | 0/4 | - | 1/4 |
| | FL.PREM | 0/2 | 0/1 | 0/1 | 0/1 | 0/2 | 0/1 |
| | FL.SFG | 0/1 | 0/1 | 0/1 | 0/1 | 0/1 | 0/1 |
| **PL** | PL.AG | 0/7 | 0/5 | 0/9 | 0/4 | 0/10 | 0/6 |
| | PL.CC | 0/8 | 0/2 | 0/9 | 0/2 | 0/10 | 0/2 |
| | PL.OPR | 0/5 | 1/8 | 1/5 | 0/8 | 1/5 | 1/8 |
| | PL.POCG | 2/3 | 0/1 | 1/3 | 0/1 | 2/3 | 0/1 |
| | PL.PREC | 0/8 | 0/10 | 0/7 | 0/10 | 0/8 | 0/10 |
| | PL.SMG | 0/20 | - | 0/20 | - | 0/21 | - |
| | PL.SPL | 0/5 | 0/3 | 0/5 | 0/3 | 0/5 | 0/3 |
| **OL** | OL.INFRAC | 0/1 | 1/1 | 1/1 | 0/1 | 1/1 | 1/1 |
| | OL.LG | 1/5 | 2/4 | 1/5 | 1/4 | 2/5 | 3/4 |
| | OL.OG | 2/10 | 0/4 | 0/10 | 1/5 | 2/10 | 1/5 |
| | OL.SUPRAC | - | 0/8 | - | 0/8 | - | 0/8 |
| **INS** | INS.ANT | 6/29 | 3/19 | 3/30 | 3/19 | 6/30 | 4/19 |
| | INS.CIR | 0/2 | 0/1 | 0/2 | 0/1 | 0/2 | 0/1 |
| | INS.POS | 0/9 | 0/13 | 1/10 | 1/13 | 1/10 | 1/13 |

**Supplementary Table 4: Significant lobe-wise hemispheric differences in HFA magnitude and activation rate.**

| TASK | LOBE | METRIC | DIRECTION | FDR CORRECTED P-VALUE |
|---|---|---|---|---|
| **ANT** | TL | Z-score magnitude | Non-Dom > Dom | 0.0397 |
| | TL | Activation Rate | Non-Dom > Dom | 0.0189 |
| | FL | Z-score magnitude | Dom > Non-Dom | 0.0498 |
| **VNT** | PL | Z-score magnitude | Non-Dom > Dom | 0.0146 |
| | FL | Z-score magnitude | Dom > Non-Dom | 0.0201 |
| **VN+ANT** | PL | Z-score magnitude | Non-Dom > Dom | 0.0276 |
| | TL | Z-score magnitude | Non-Dom > Dom | 0.0126 |

Abbreviations: TL=temporal lobe, FL=frontal lobe, PL=parietal lobe

**Supplementary Table 5: Channel wise diagnostic performance of HFA activation in predicting DCS status, at the stringent threshold of z-score >0.8.** Values are presented as estimate (95% Confidence Interval).

| TASK | TP | FN | FP | TN | SENS | SPEC | PPV | NPV | BA |
|---|---|---|---|---|---|---|---|---|---|
| **VNT** | 8 | 37 | 42 | 557 | 0.18 (0.09-0.31) | 0.93 (0.91-0.95) | 0.13 (0.08-0.29) | 0.95 (0.92-0.95) | 0.55 |
| **ANT** | 4 | 41 | 40 | 559 | 0.09 (0.04-0.21) | 0.93 (0.91-0.95) | 0.07 (0.04-0.21) | 0.94 (0.91-0.95) | 0.51 |
| **VN & ANT** | 11 | 34 | 69 | 530 | 0.24 (0.14-0.39) | 0.88 (0.86-0.91) | 0.14 (0.08-0.23) | 0.94 (0.92-0.96) | 0.56 |

Abbreviations: TP = true positives, FN = false negatives, FP = false positives, TN = true negatives, Sens = sensitivity, Spec = specificity, PPV = positive predictive value, NPV = negative predictive value, BA = balanced accuracy.

**Supplementary Table 6: Channel wise diagnostic performance of HFA activation in predicting DCS status, at the permissive threshold of Z-score >0.3.** Values are presented as estimate (95% Confidence Interval).

| TASK | TP | FN | FP | TN | SENS | SPEC | PPV | NPV | BA |
|---|---|---|---|---|---|---|---|---|---|
| **VNT** | 16 | 29 | 133 | 542 | 0.36 (0.23-0.5) | 0.78 (0.74-0.81) | 0.09 (0.07-0.17) | 0.95 (0.92-0.96) | 0.57 |
| **ANT** | 15 | 30 | 115 | 484 | 0.33 (0.21-0.48) | 0.81 (0.77-0.84) | 0.1 (0.07-0.18) | 0.95 (0.92-0.96) | 0.57 |
| **VN & ANT** | 25 | 20 | 190 | 409 | 0.56 (0.14-0.69) | 0.66 (0.64-0.72) | 0.09 (0.08-0.17) | 0.96 (0.93-0.97) | 0.62 |

See supp. table 5 for abbreviations.

**Supplementary Table 7: Region wise diagnostic performance of HFA activation in predicting DCS status, at the stringent threshold of z-score >0.8.** Values are presented as estimate (95% Confidence Interval).

| TASK | TP | FN | FP | TN | SENS | SPEC | PPV | NPV | BA |
|---|---|---|---|---|---|---|---|---|---|
| **VNT** | 10 | 18 | 31 | 184 | 0.36 (0.21–0.54) | 0.86 (0.80–0.90) | 0.24 (0.14–0.39) | 0.91 (0.86–0.94) | 0.61 |
| **ANT** | 4 | 23 | 32 | 177 | 0.14 (0.06-0.32) | 0.84 (0.79-0.89) | 0.09 (0.04-0.25) | 0.9 (0.83-0.92) | 0.49 |
| **ANT+VN** | 12 | 16 | 51 | 172 | 0.43 (0.27–0.61) | 0.77 (0.71–0.82) | 0.17 (0.11–0.30) | 0.92 (0.87–0.95) | 0.60 |

See supp. table 5 for abbreviations.

**Supplementary Table 8: Region wise diagnostic performance of HFA activation in predicting DCS status, at the permissive threshold of Z-score >0.3.** Values are presented as estimate (95% Confidence Interval).

| TASK | TP | FN | FP | TN | SENS | SPEC | PPV | NPV | BA |
|---|---|---|---|---|---|---|---|---|---|
| **VNT** | 14 | 14 | 89 | 126 | 0.50 (0.33–0.67) | 0.59 (0.52–0.65) | 0.14 (0.08–0.22) | 0.90 (0.84–0.94) | 0.57 |
| **ANT** | 14 | 13 | 76 | 133 | 0.52 (0.34–0.69) | 0.64 (0.57–0.70) | 0.16 (0.09–0.24) | 0.91 (0.85–0.95) | 0.57 |
| **ANT+VN** | 21 | 8 | 116 | 107 | 0.75 (0.53–0.85) | 0.45 (0.42–0.55) | 0.13 (0.10–0.22) | 0.94 (0.87–0.96) | 0.6 |

See supp. table 5 for abbreviations.
